# Use of an integrated pan-cancer oncology enrichment NGS assay to measure tumour mutational burden and detect clinically actionable variants

**DOI:** 10.1101/2020.02.01.20019992

**Authors:** Valerie Pestinger, Matthew Smith, Toju Sillo, John M Findlay, Jean-Francois Laes, Gerald Martin, Gary Middleton, Phillipe Taniere, Andrew D Beggs

**Author notes:** Correspondence to: Andrew D Beggs, Surgical Research Laboratory, Institute of Cancer & Genomic Science, University of Birmingham, Vincent Drive, Birmingham, B15 2TT.

## Abstract

**Introduction:** The identification of tumour mutational burden (TMB) as a biomarker of response to PD-1 immunotherapy has necessitated the development of genomic assays to measure this. We carried out comprehensive molecular profiling of cancers using the Illumina TruSight Oncology panel (TSO500) and compared to whole genome sequencing.

**Methods:** Cancer samples derived from formalin fixed material were profiled on the TSO500 panel, sequenced on an Illumina NextSeq 500 instrument and processed through the TSO500 Docker Pipeline. Either FASTQ files (PierianDx) or VCF files (OncoKDM) were processed to understand clinical actionability

**Results:** In total, 108 samples (a mixture of colorectal, lung, oesophageal and control samples) were processed via the DNA panel. There was good correlation between TMB, SNV, indels and CNV as predicted by TSO500 and WGS (R2>0.9) and good reproducibility, with less than 5% variability between repeated controls. For the RNA panel, 13 samples were processed, with all known fusions observed via orthogonal techniques detected. For clinical actionability 72 Tier 1 variants and 297 Tier 2 variants were identified with clinical trials identified for all patients.

**Conclusions:** The TruSight Oncology 500 assay accurately measures TMB, MSI, single nucleotide variants, indels, copy number/structural variation and gene fusions when compared to whole genome sequencing and orthogonal technologies. Coupled with a clinical annotation pipeline this provides a powerful methodology for identification of clinically actionable variants.

## Introduction

Recent developments in next-generation sequencing and tumour immunology have allowed the discovery that targeting the CTLA4, PD-1 and PD-L1 receptors using therapeutic monoclonal antibodies (1) can unmask cancer to the immune system, facilitating its immune-mediated destruction. Although initial trials of PD-1 inhibitors had mixed results (2, 3), as with previous targeted therapies, it was determined that a specific tumour genotype was required in order for these inhibitors to be effective, leading to the finding that dramatic regression of tumours could occur with the correct genotype.

In order for tumours to become immunogenic, a high neoepitope load must be generated via hypermutation (4-6), ideally indel/frameshifts or non-synonymous mutations that generate novel proteins that can be recognised by the immune system. These neoepitopes can then be presented via MHC in order to aid immune killing (7).

The CHECKMATE (8-10) series of trials have suggest that a specific threshold of “tumour mutational burden” (TMB) must be reached in order for PD-1 blockade to become effective. Although TMB has variable definitions, it is broadly accepted (9) as the number of missense mutations in the tumour genome, either divided by the size of the exome panel (35-45 Mb) or via the size of the human genome for whole genome sequencing (3.3 Gb). Based on the CHECKMATE trials, the suggested TMB threshold is greater than 10 mutations/mb, based on the objective response rates of the tumours in these studies not improving much beyond this threshold.

Initially, TMB was measured using whole genome and whole exome sequencing (11), however these technologies are not cost effective currently for routine use in the clinic. Despite the falling cost of next-generation sequencing reagents, the volume of data required for sufficient coverage of either whole genome (200-300 Gb for 60X read depth) or whole exome (4-5 Gb for 100X read depth) sequencing make it impractical except for dedicated sequencing cores. Secondly, even with high read depth, sufficiently deep coverage in order to identify rare subclonal (12) mutations that may contribute to the neoantigen load is required, of the order of 500X. Thus, whole genome/exome coverage is not cost effective (13).

Rizvi et al (13) demonstrated that in order to accurately measure TMB using a NGS-based assay, a panel size of at least 1.5 megabases is required. This panel size offers opportunities for a pan-cancer assay, as a panel of this size could cover the majority of known driver genes across multiple cancer types. In designing an oncology assay, ideally other types of variations would be included. Recent studies (14, 15) have shown the potential utility of selecting targeted therapies using large gene panels and therefore a panel should include mutations associated with targeting therapies.

An additional advantage of panel-based designs is the ability to enrich RNA targets. Recent studies have shown the importance of RNA fusions such as the TMPRSS-ERG fusion in prostate cancer (16), the FGFR2 fusion in cholangiocarcinoma (17), and the NTRK fusion in lung and other cancers (18). These fusions are either targetable with molecularly targeted agents (e.g. larotectinib (19) or pemigatinib (20)) or are prognostically relevant (i.e.

TMPRSS-ERG).

An ideal oncology panel-based assay would have several characteristics (21): enrichment chemistry rather than PCR chemistry for identification of rare alleles with straightforward library preparation; a broad panel that targets the majority of DNA & RNA alterations in cancer; rapid run time; prediction of novel biomarkers such as TMB; and a standardised, reproducible analysis pipeline that can be used in a clinical setting.

In this study, we present our initial results using the Illumina TruSight Oncology 500 assay across a range of cancer types. We benchmarked it against whole genome and exome sequencing, as well as determining its ability to detect RNA fusions and copy number variants.

## MATERIALS AND METHODS

### Patient samples & ethics

Patient samples were from three cohorts – colorectal, oesophageal, and lung. For the colorectal cohort, fresh frozen tumours were obtained from an internal cohort of patients that had undergone whole genome sequencing as part of a pre-pilot prior to the introduction of the 100,000 Genomes project. Matched FFPE blocks from the tumour were obtained and used for the assay. For the oesophageal cohort, sequential oesophageal cancers underwent whole exome sequencing from a cohort collected at the Oxford Cancer Centre. For the lung cohort, patient samples were obtained from those undergoing routine testing for EGFR mutation status at the Molecular Pathology department of the University Hospitals Birmingham NHS Foundation Trust.

Ethics for the study was obtained from the Oxford Ethics committee (reference 05/Q1605/66)

### Nucleic acid extractions and quality assessment

DNA and RNA were extracted from 2 × 5 µm FFPE scrolls on the Covaris E220 evolution (520220, Covaris Ltd, Woodingdean, Brighton, UK) using the truXTRAC FFPE total NA Kit - Column Purification (520220, Covaris Ltd, Woodingdean, Brighton, UK) following manufacturer’s protocol.

65% isopropanol was used during RNA purification. On-column DNA digestion was performed after the first wash during RNA purification using the TURBO DNA-free kit (AM1907, Invitrogen, ThermoFisher Scientific, Paisley, UK) following Covaris protocol. DNA and RNA concentrations were measured on the Qubit 3 Fluorometer (ThermoScientific, Paisley, UK) and DV200 (he percentage of fragments >200 nucleotides in size) was assessed using Tapestation 2200 (Agilent, Cheshire, UK). DNA quality was determined by the Infinium HD FFPE QC Assay Protocol (15020981, Illumina, Cambridge, UK).

RNA samples with a DV200 ≥30% and DNA samples with a Delta Cq value ≤5 were used for downstream applications.

### Library preparation

DNA libraries were prepared using the hybrid capture-based TruSight Oncology 500 Library Preparation Kit (Illumina, San Diego, CA, USA) following Illumina’s TruSight Oncology 500 Reference Guide (Document # 1000000067621 v00, Illumina Cambridge, UK) with the following modifications.

gDNA was sheared using the Covaris E220 evolution (Covaris Ltd, Woodingdean, Brighton, UK), 8 micro TUBE – 50 AFA Fiber Strip V2 (520174, Covaris Ltd, Woodingdean, Brighton, UK) and Rack E220e 8 microTUBE Strip V2 (500437, Covaris Ltd, Woodingdean, Brighton, UK). Size of dsDNA fragments (90-250bp) was confirmed using Tapestation 2200 (Agilent, Cheshire, UK) after shearing. A HorizonDx HD753 control (Horizon Discovery, Cambridge, UK) was included with every set of 7 test samples. When no beads were involved, reagents were mixed by pipetting up and down 10 times. Before the bead based normalization, libraries were quantified and sized on the Qubit 3 Fluorometer (ThermoScientific, Paisley, UK) and Tapestation 2200 (Agilent, Cheshire, UK), respectively.

10 µl of each normalized DNA library (max of 8 libraries per pool) were pooled and incubated at 96°C for 2 min. The tube containing the library pool was immediately inverted two times to mix, centrifuged briefly, and placed on ice for 5 min. 10 µl of the library pool were mixed with 190 µl HT1 to make a 1:20 dilution (DIL1). 40 µl of DIL1 were mixed with 1360 µl HT1 (for a final library concentration of 1.5 pM) and 2.5 µl of denatured 20 pM PhiX added (1%). Libraries were sequenced on an Illumina NextSeq 500 instrument.

For whole genome sequencing libraries, 1 ug of DNA was prepared using the TruSeq DNA library preparation kit (Illumina, San Diego, CA, USA) and sequenced across 4 lanes of a HiSeq 2500 (Illumina, San Diego, CA, USA).

## Bioinformatics

Raw sequencing output was transferred from the sequencing instrument to a bioinformatics server running Ubuntu 18.04LTS. A pre-supplied Docker image (The TruSight Oncology 500 pipeline; Illumina, San Diego, CA, USA) was used to generate TMB and Microsatellite Instability (MSI) calls. The pipeline consists of several steps. Initially, raw bcl files are converted to sample-specific FASTQ files as specified by the sample index. FASTQ files were then aligned against the hg19 reference genome using Isaac 4, local realignment to indels was performed, paired-end reads were stitched together, followed by variant calling with the somatic sample caller Pisces. Germline variants were filtered using a proprietary database, then the called variants were annotated to identify synonymous and non-synonymous variants. Actual coverage of the panel compared to the reference coverage was computed and TMB was calculated based on the number of synonymous and non-synonymous mutations detected divided by the size of the panel successfully sequenced.

Small variants were exported from the TSO500 pipeline and annotated using VEP, then converted using vcf2maf and imported into the maftools module of R/Bioconductor.

TMB calls for whole genome sequenced control data were carried out using the Genomics England v3 pipeline for calling tumour-normal pairs and used to compare to calls from the TSO-500 pipeline. In brief, this pipeline utilised Isaac v3 to align sequence data to the hg19 genome, followed by copy number variant calling using Canvas and structural variant calling using Manta. CNV calls for the TSO500 files were obtained using the Craft copy number caller set in somatic tumour only mode. Overlaps were computed using bedtools. SV calls for the TSO500 files were obtained using the Manta structural variant caller set in tumour only mode with a custom modification to the C++ code of the Manta SV caller to enable detection with less read support and on amplicon sequencing data. SV overlaps were computed using bedtools.

For clinical actionability, raw FASTQ files (CGW, PierianDx, St. Louis, MO, USA) and UMI collapsed VCF files obtained from the TSO500 v1 Docker image (OncoKDM, OncoDNA, Gosselies, Belgium) were uploaded to their respective data portals and run in their standard analysis mode. The Clinical Genomic Workspace (CGW; PierianDx, St Louis, MO, USA) is a secure web-based HIPAA- and GDPR-compliant platform for clinical decision support management. Initially developed by one of the very first medical institutes to launch a routine clinical next generation sequencing service for cancer and complex inherited diseases, the CGW encompasses a rules engine built on a curated knowledgebase that is updated weekly. Information from over 18 millions publications, including FDA and EMA approvals, NCCN, AMP and ESMO guidelines and pubmed articles is coupled with public data sources such as population databases, dbSNP, TCGA, ClinVar and COSMIC in order to annotate and pre-classify variants for interpretation. Uniquely, the CGW utilises the World’s largest clinical interpretation sharing network that provides variant interpretations in the context of the specific disease defined for the patient at time of accessioning. Although no patient data is transferred, network members can view the clinical interpretations supplied to the clinical team of the provider institution (giving the most up-to-date information with true clinical provenance). Actionability calls were downloaded according to standard Association of Molecular Pathology (AMP) tiers.

OncoKDM is a secure web-based ISO27001, IS013485 and GDPR-compliant platform for clinical decision support management and clinical report sharing. Initially developed for their proprietary OncoDEEP products that have been on the market since 2013, OncoKDM encompasses a proprietary daily/weekly curated knowledge database of 22,000 genes, 3,886,000 variants, 792 drugs (including FDA and EMA approvals, NCCN, Compermed and ESMO guidelines), 5000 associated clinical trials and 7000 associated publications. Coupled with several public data sources, OncoKDM accurately retrieves the biological and clinical information for proper data interpretation and has already been used for six years thanks to the sharing platform OncoSHARE used by 6,500 healthcare professionals in 50 countries worldwide.

## Results

### DNA – quality metrics

In total, 108 samples were profiled using the assay, with a median sample age of 2 years (range 4 months-10 years). All samples were from FFPE blocks. This input for all assays was 40 ng of DNA and 40 ng of RNA. Suitability of samples for sequencing was determined using the real time PCR-based Illumina FFPE QC assay. For a sample to pass initial QC, the dCt must be under 5. Study samples had a median dCt of 2.46 (IQR 1.73-4.3). The maximum dCt run was 13.57, and all samples bar one passed TMB calling. Eight samples failed MSI calling due to poor sample quality (dCt=5-13.57). Samples were all sequenced despite the initial QC metrics to determine the validity of this measure in determining samples for sequencing.

In terms of DNA sequencing metrics, the median insert size was 92.5 bp (IQR 80-112 bp), the median exon coverage was 185x (IQR 123-247x), and 98.1% and 90.5% of all samples were covered at least 50x and 100x respectively. The median reads per sample was 126M (IQR 105M-138M reads); there was a median of 0.9% (IQR 0.25-2.7%) chimeric reads per sample. Median read enrichment was 82.1% (IQR 79-85%).

### Mutational coverage & spectrum

In all 108 samples mutational coverage of the panel was successfully performed with little probe drop out. Figures 1, 2, and 3 show the variant classification results. There was a median of 14 variants per sample (range 2-479). The predominant mutation type was a missense SNP, followed by frame shift deletion. The top ten most commonly mutated genes were TP53 (73%), APC (54%), FLT3 (50%), LRP1B (27%), SPTA1 (20%), BRCA2 (20%), KRAS (30%), PIK3CA (22%), ARID1A (21%), and CREBBP (20%). Within the sample subgroups, colorectal cancer was significantly enriched for APC mutations (40/54 CRC samples), ZFHX3 (13/54 samples) and FBXW7 (13/54). Oesophageal cancer was significantly enriched for EPHA7 mutations (3/9 sample), and TP53 (9/9 samples). For lung, RBM10 (5/22 samples) and MGA (7/22 samples) were significantly enriched.

**Figure 1:**
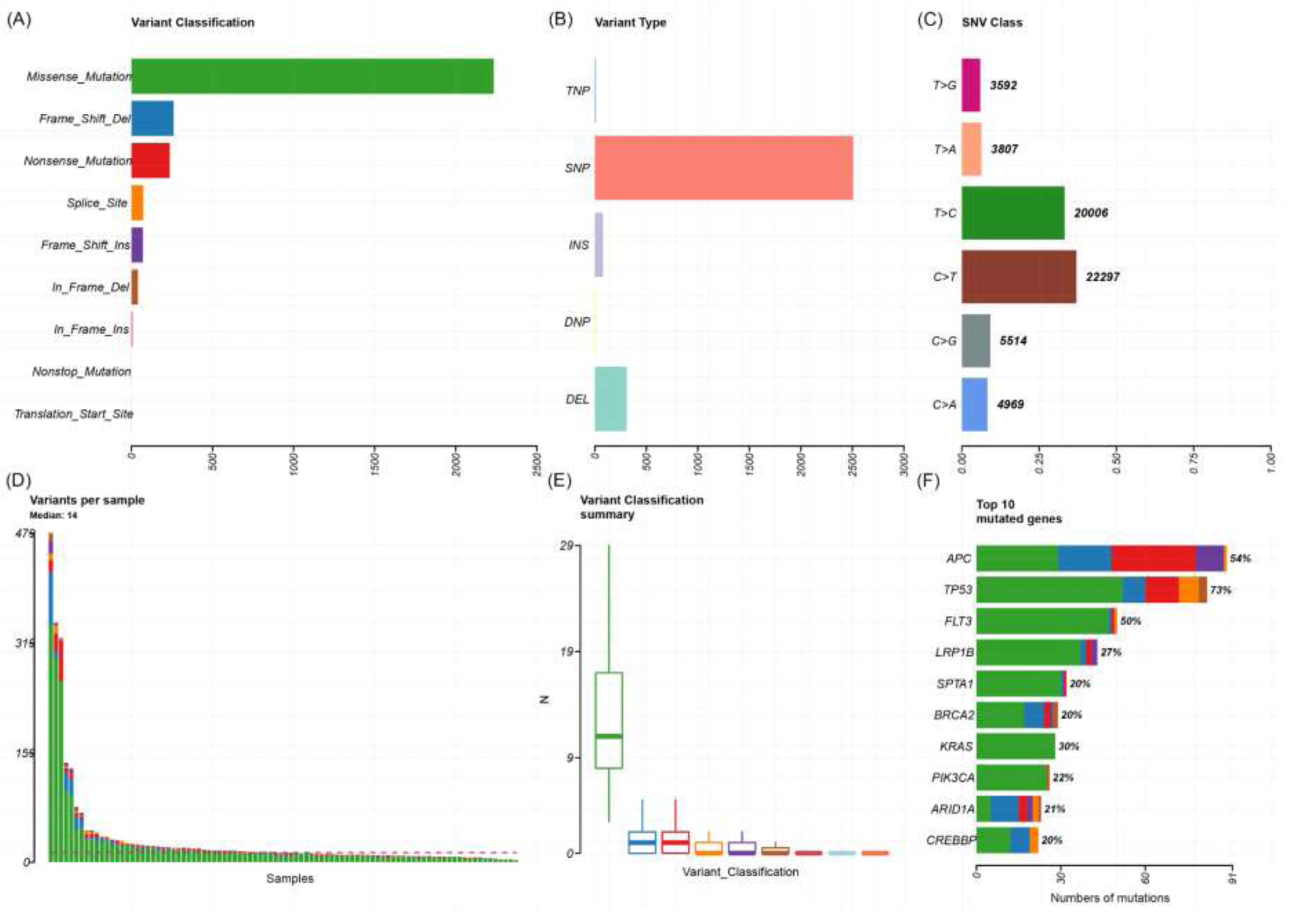
Plot of variant classification for all samples using TSO500. A, Total number of variants detected by variant classification. B, Total number of variants detected by variant type. C, Total number of variants detected by SNV class. D, Number of variants per sample. E, Box plot of the number of variants within each classification per sample. F, Top ten mutated genes. For D, E, and F, the colours are equivalent to A. TNP, trinucleotide polymorphism; SNP, single nucleotide polymorphism; INS, insertion: DNP, dinucleotide polymorphism; DEL, deletion.

**Figure 2:**
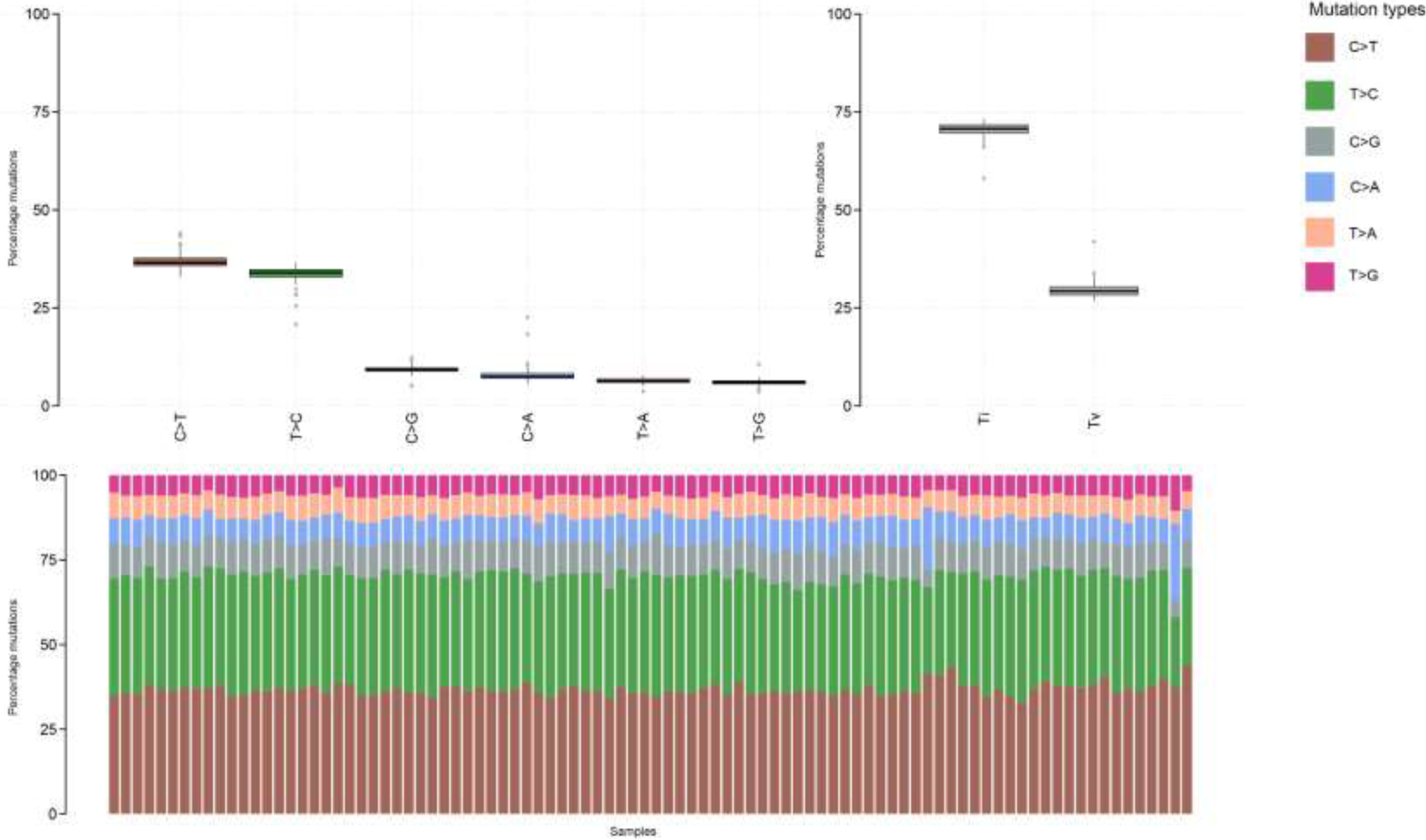
Mutational plot for all samples in TSO500 assay. Top left panel = type of mutation (frequency in percent), top right panel = Transitions (Ti) vs. Transversion (Tv). Bottom panel = proportion of mutations.

**Figure 3:**
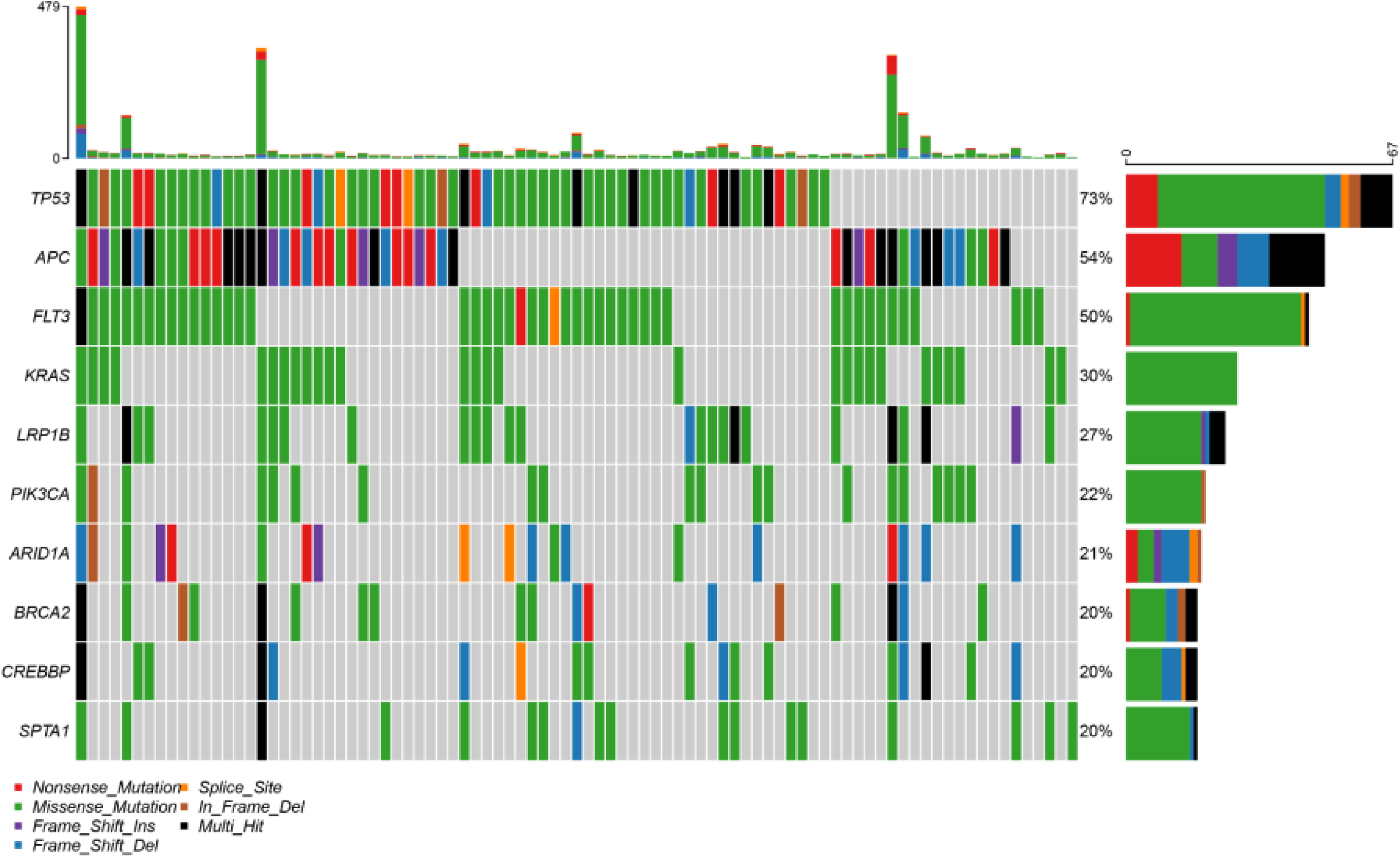
OncoStrip plot of common variants across samples. Genes on Y-axis; Samples on Axis.

In terms of mutational spectrum, a predominance of C>T transversions was seen, as the samples were all derived (with the exception of controls) from FFPE tissue blocks. The predominant mutational signatures (22) seen in the samples were signature 10 (defects in polymerase POLE), signature 5 (due to tobacco smoke), and signature 6 (defective mismatch repair), which fit well with the source of the samples (lung, colorectal and oesophageal) as well as the fact that several hypermutant samples were deliberately chosen for the project.

### Precision of control calls

In order to understand the ability of the assay to detect low variant allele frequencies (VAF), assessment of the VAF was performed for two known low VAF mutations in the HD753 cell line (Figure 4). This cell line has validated mutations in *AKT1* (E17K, chr14: 105246551C>T) with a VAF of 0.05 and in *PIK3CA* (E545K, chr3: 178936091G>A) with a VAF of 0.056. The same control was run across 12 runs, with *AKT1* median VAF=0.059 (IQR 0.037-0.072) and *PIK3CA* median VAF=0.036 (IQR 0.033-0.0493).

**Figure 4:**
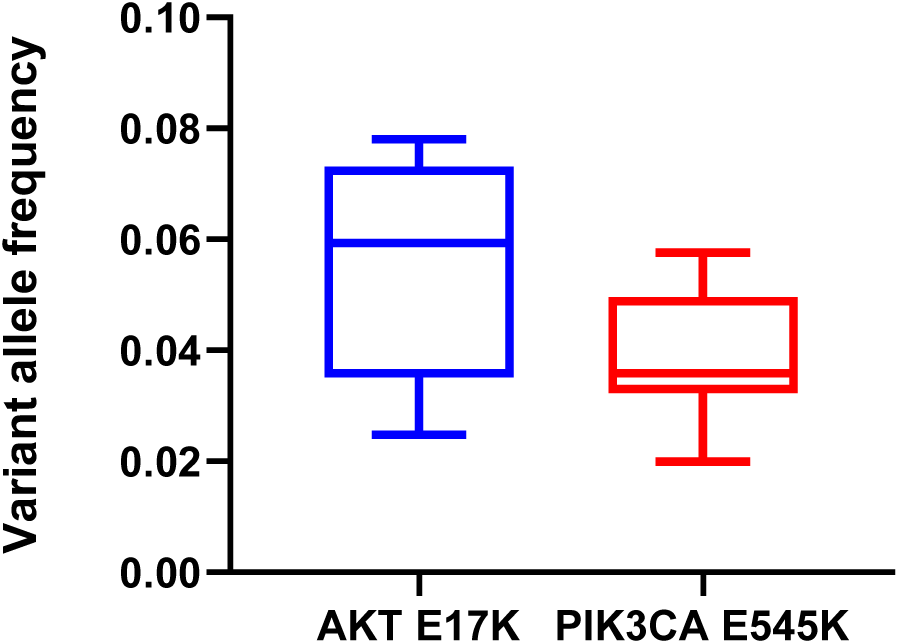
Box and whisker plot of repeated measures of variant allele frequency of known low VAF variants in a HD753 control. Known VAF is 0.054 for the AKT E17K mutation and 0.05 for the PIK3CA E545K mutation.

### Copy number calls

A subset of 24 sample underwent copy number calling with the Craft pipeline. A variety of copy number gains and losses were detected in the 520 genes profiled on the TSO500 panel. The HD753 control was used to determine whether the observed copy number calls (Figure 5) correlated with known copy number changes: amplifications in *MET* (CNV=4.5) and *MYC-N* (CNV=9.5). The amplification in *MET* was observed in all control samples with an average CN=4 and in *MYC-N* with an average CN=9.1. No whole gene deletions were present in the control samples, but were observed in a variety of samples in the tumour cohort.

**Figure 5:**
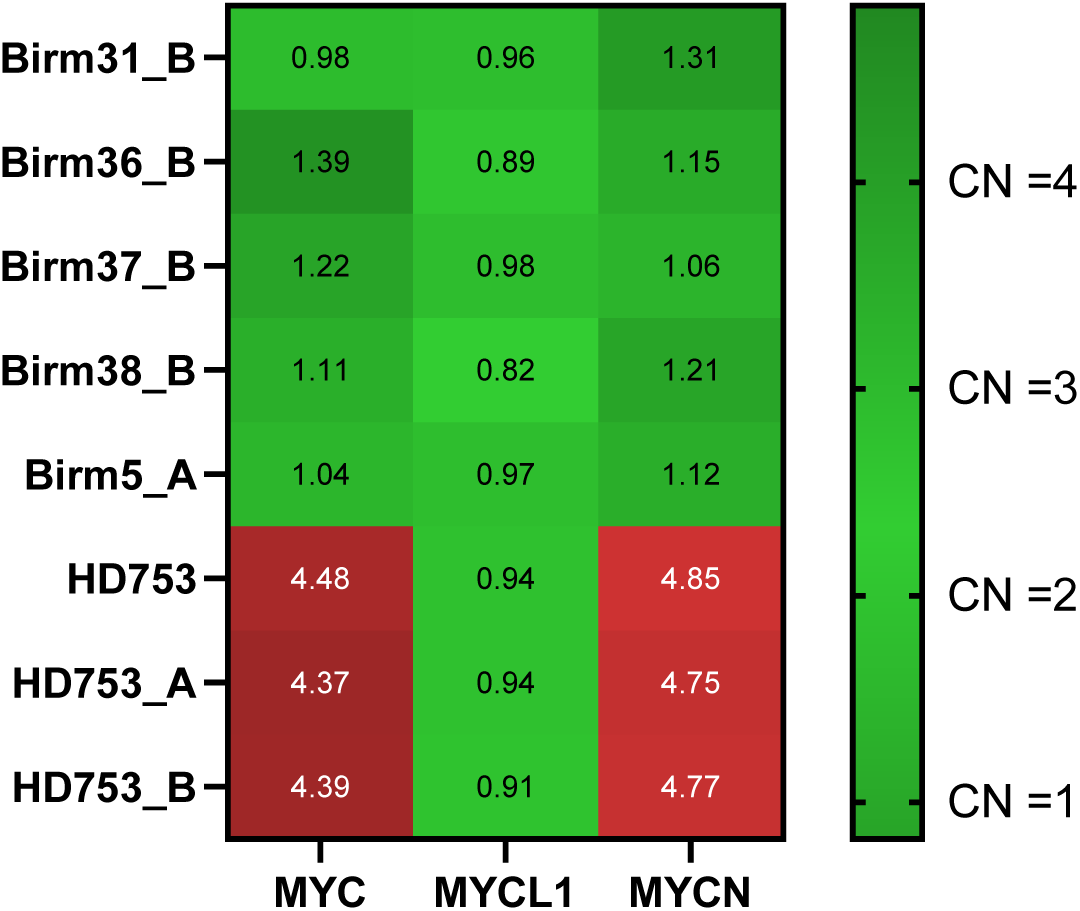
Copy number heatmap of detected copy number in HD753 samples with known amplification in MYCN and MYC. Sample IDs on Y-axis.

### Structural variant calls

The HD753 control is known to have a variety of structural variants including a *SLC34A2*/*ROS* fusion (VAF = 5.6%) and *CCDC6*/*RET* fusion (VAF = 5.0%). With use of a custom pipeline there was evidence for detection of both fusions: 7/506 reads supported the SLC34A2/ROS fusion and 5/498 reads supported the CCDC6/RET fusion. A variety of structural variants were observed in the tumour cohort. In addition, long indels were successfully detected by the Manta pipeline, specifically a 14bp deletion in *EGFR* (NM_005228.5:c.2235_2249del) known to be present in the HD753 control.

### Tumour mutational burden (TMB) & Microsatellite instability (MSI)

TMB calling was successfully performed in 107/108 samples. The one failure was a very poor sample quality that failed hybridisation. There was good correlation between TMB determined by TSO500 and WGS (R^2^=0.9, Figure 6). The median TMB was 8.6 mutations(mut)/Mb (range 0.85-325 mut/Mb, Figure 7). Several known hypermutant tumor samples were deliberately run first, including a somatic POLE mutant colorectal cancer (reported TMB 261.71 mut/Mb in WGS sample), a somatic MLH1 mutant colorectal cancer (reported TMB 67.43 mut/Mb) and a somatic MSH6 mutant colorectal cancer (reported TMB 104.0 mut/Mb). As the HD753 control was run in each experiment, we compared the reproducibility of the TMB measurement for this control sample. There was a median TMB of 311 muts/mb (range 289-325) in the HD753 control, a variance in TMB score of +/- 5%. Comparison to The Cancer Genome Atlas (TCGA) tumour cohorts was performed in mafTools and is shown in Figure 8.

**Figure 6:**
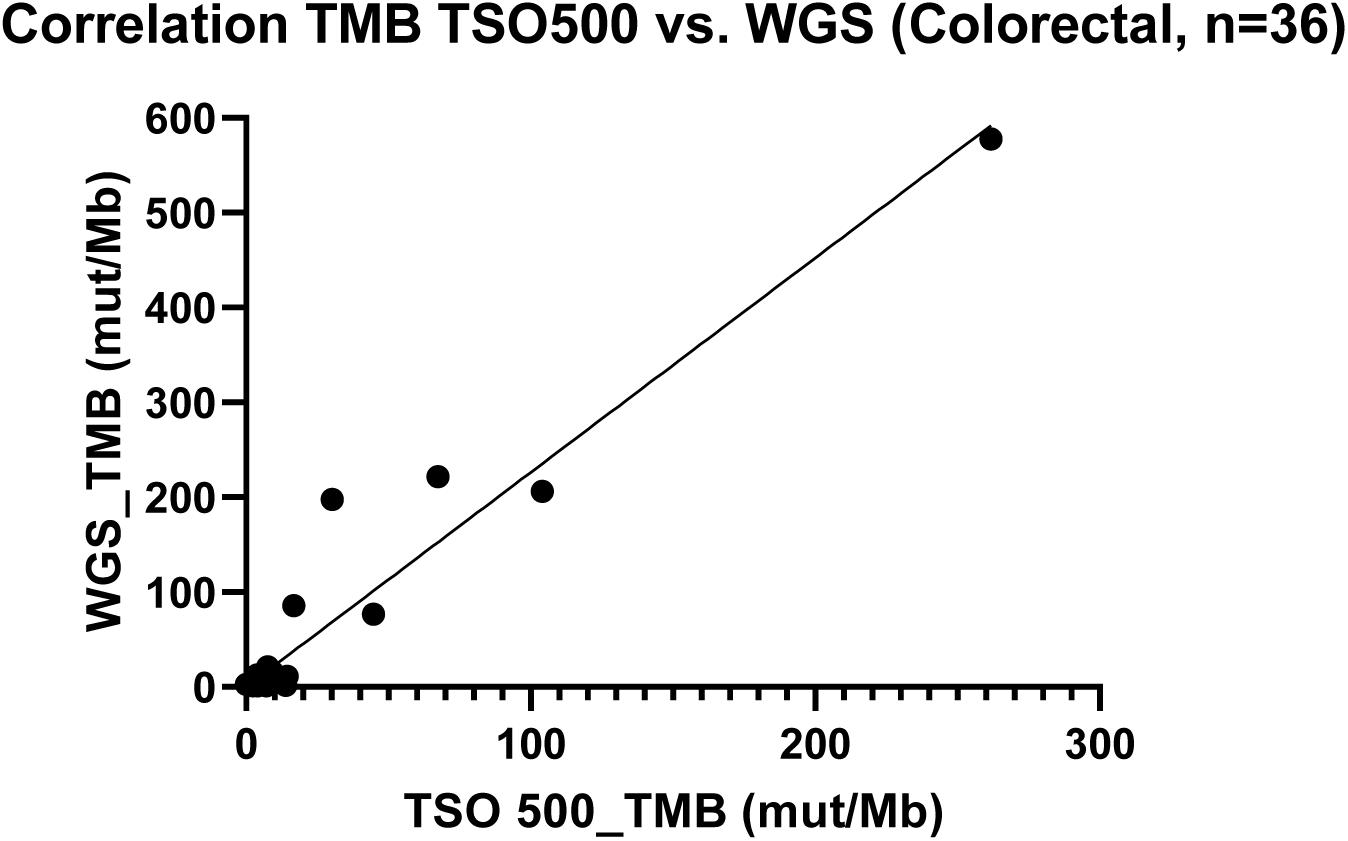
Correlation between tumour mutational burden as measured by TSO500 assay vs. PCR-free whole-genome sequencing on colorectal samples (R^2^=0.98, p<0.001)

**Figure 7:**
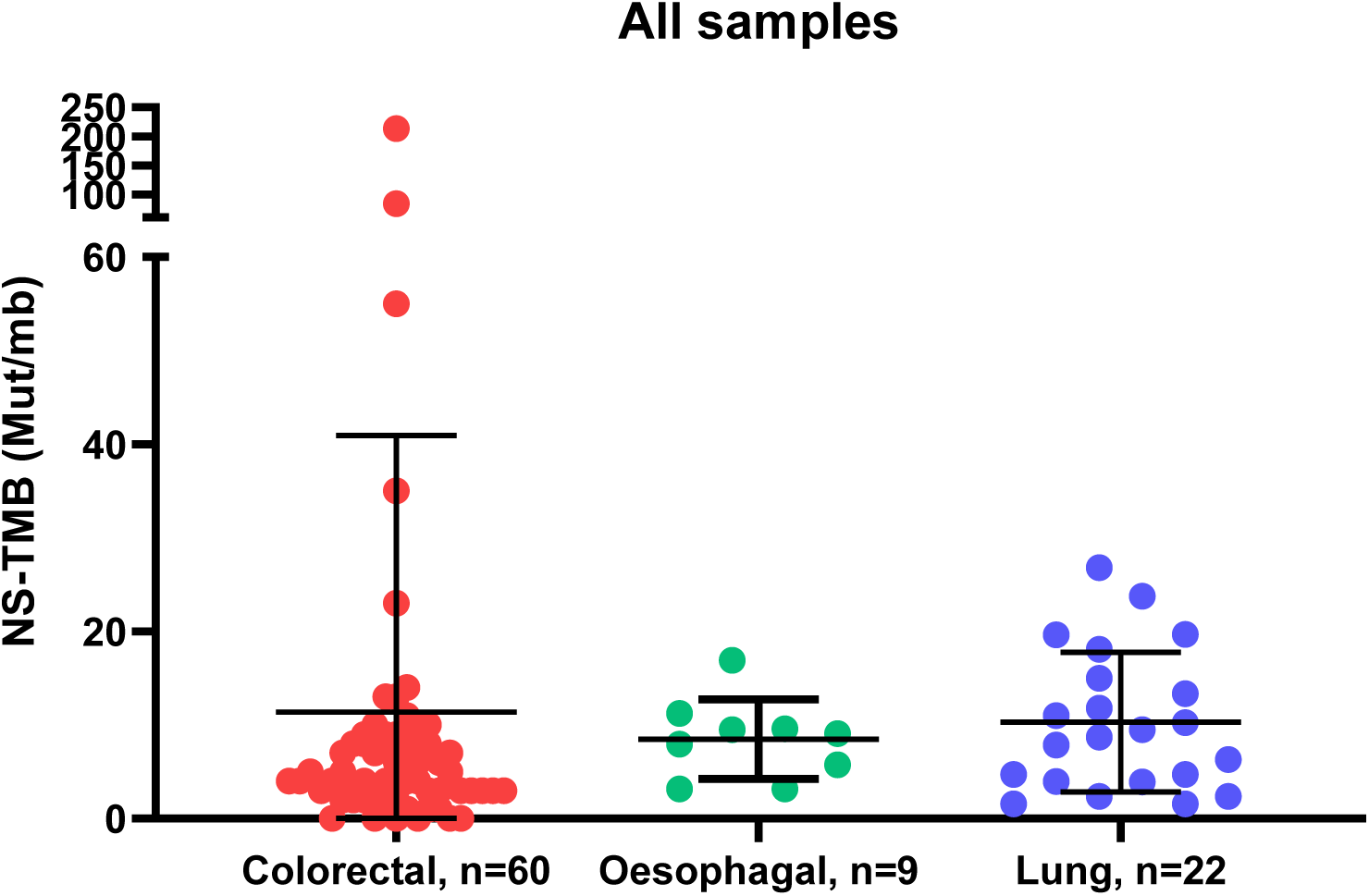
Distribution of mutations/megabase of DNA by tumour type. Y-axis = Non-synonymous tumour mutational burden in mutations per megabase of DNA.

**Figure 8:**
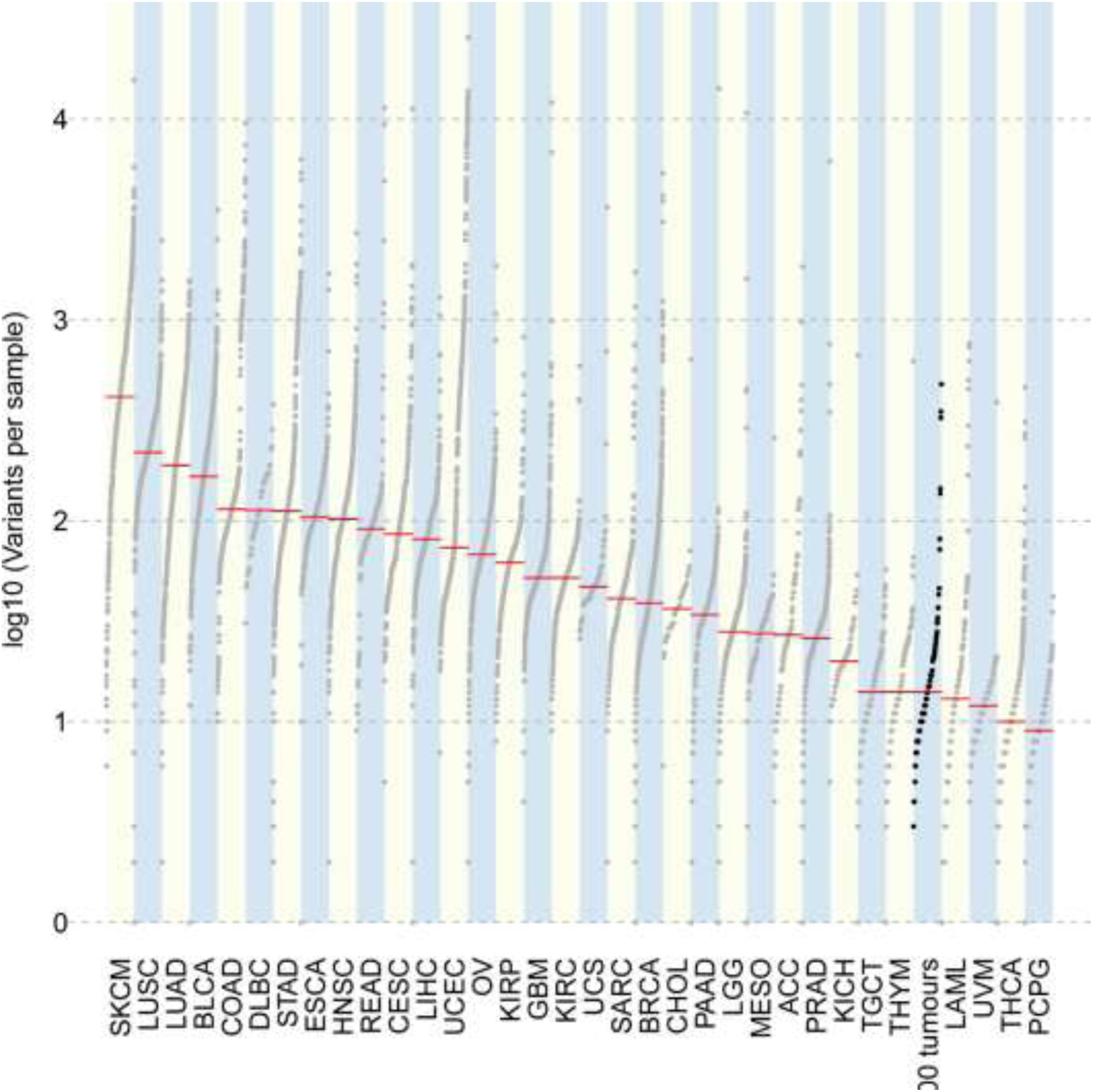
Plot of mutational burden in TSO500 trial tumour set as compared to tumours from TCGA Atlas. TSO500 trial tumour set shown in black. Y-axis = log10(Variants per sample).

For microsatellite instability (Figure 9), the threshold for classification as MSI-H was >10% of microsatellite sites being unstable. Using this threshold, both known somatic mismatch repair mutant (MLH1 and MSH6) cancers were MSI-H with 55% and 67% of sites being unstable respectively. Reassuringly, the POLE mutant cancer had 2% of MSI sites being unstable, meaning it was microsatellite stable (MSS) as is typical in POLE mutant cancer.

**Figure 9:**
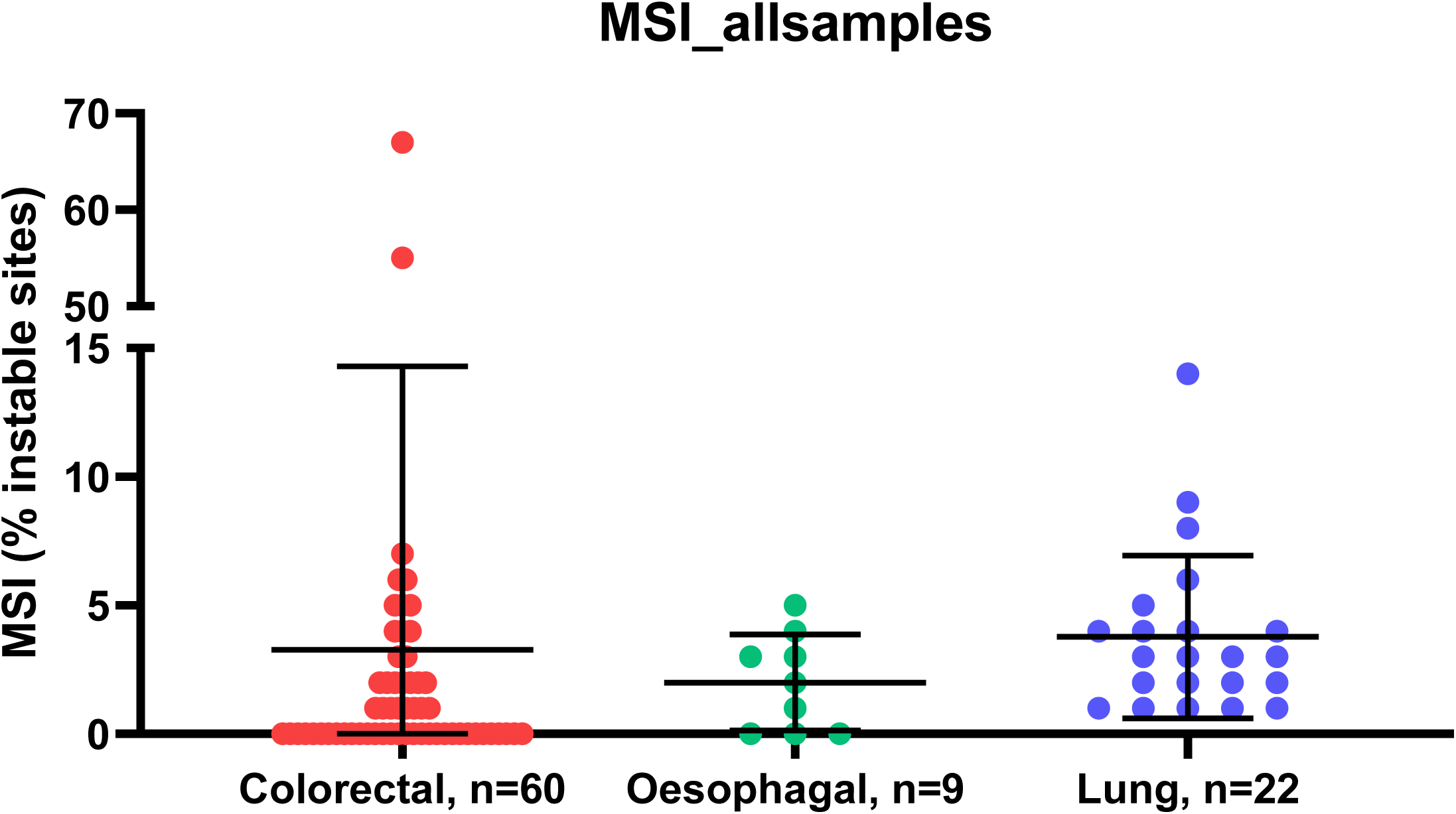
Percentage of microsatellite unstable probes against tumour type. Colorectal = red, oesophageal = green, lung = blue.

### RNA fusions

RNA fusion analysis was carried out on 13 samples, of which 6 had known fusions. Fusions were detected between ETV6/NTRK3 (3 samples), RBPMS/NTRK3 (1 sample), EML4/ALK (1 sample), and TG/RET fusion (1 sample). All fusions that had previously been identified by FISH were detected using this methodology. A fusion was detected in one sample between ETV6/NTRK3 that had not been identified via FISH, however the fusion was supported by 12,627 reads in the sequencing run, which we felt was unlikely to be a false positive and therefore labelled it as a true fusion.

### Clinical actionability

In order to recover as many clinically actionable variants as possible, mutational calls were fed into the OncoDNA OncoKDM (Gosselies, Belgium) and PierianDx CGW (St. Louis, MO, USA) pipelines. OncoDNA was provided with the VCFs coming from the Illumina pipeline and PierianDX started the analysis directly from the FASTQ files. Also, in order to take an overview of pathway mutations and potential targets, the *OncogenicPathways* function of *mafTools* was used to generate a list of druggable pathways. The RAS pathway had 39/85 (45.9%) genes mutated, the PI3K pathway 20/29 genes (70.0%), and TGF-beta 6/7 (85.7%) genes mutated.

Using this combined approach, more than 72 Tier 1 variants and more than 297 Tier 2 variants were identified between the two pipelines. Twenty-one samples were classified as TMB High, 19 as TMB medium and the remainder (64) as TMB-low. Clinical trials could also be identified for all samples, with a median of 22 (Range 4-105) trials suggested per sample. The most common actionable mutations observed were BRAF p.V600E (18 samples), KRAS p.G13D (14 samples) and KRAS p.G12D (12 samples). For other tiers, there were 8,175 tier 3 mutations and 17,649 Tier 4 mutations detected.

PierianDx and OncoKDM pipelines were not directly comparable in this study because of the differing inputs (FASTQ for PierianDx, VCF for OncoKDM) but the combination of both platforms provided a comprehensive variant overview.

## Conclusions

We utilised the Trusight Oncology 500 assay in order to understand its utility and accuracy in determining both the tumour mutational burden and druggable mutation calls in cancer. One of the key challenges with patient testing (23) is the ability to take a patient biopsy sample, with limited input material and produce sequencing data and mutational calls of sufficient quality in order to make decisions on target selection and drug therapy (24).

The assay was designed in its first iteration to measure tumour mutational burden as a surrogate marker for response to anti-PD1 immunotherapy as multiple studies have shown a correlation between TMB and response to this type of therapy (8, 13). The TSO500 assay performs well in this respect with accurate measurement of TMB when compared to whole genome sequencing. Taking a threshold of 10 mut/mb as “TMB-high” (i.e. that which would have benefit for immunotherapy), we found that the TSO500 assay was able to classify samples with 100% accuracy. The precision of the calls varies at the extremes of TMB value, undoubtedly as a factor of panel size in calling TMB at extremely high levels. We conclude that the TSO500 pipeline is usable in clinical determination of TMB status across a range of clinical sample types and DNA inputs.

We successfully detected microsatellite instability in all samples that were known to be MSI-H using TSO500. MSI detection using NGS has been shown to be feasible (25) previously using a variety of software solutions, usually relying on off-target reads (26), but other assays have used dedicated MSI probes (like the TSO500). We have found that the performance of this approach is variable, as the probes are vulnerable to drop out in FFPE samples. We propose that TMB instead may be a good surrogate biomarker for MSI, as a range of 30-80 mut/mb is typically seen in MSI tumours as opposed to MSS POLE/POLD1 tumours which typically have greater than 150 mut/mb.

A key requirement for clinical specimens is the ability to process low-input specimens as well as the ability to detect the low variant allele frequencies (VAF) associated with these specimens (27). Reassuringly, we found that the TSO500 assay performed well at its recommended input concentration and also below these levels. Within our control samples with known VAF (of approx. VAF=0.05), we determined that there was good precision and reproducibility with minimal variability. Another advantage to tolerance of low sample input is the possibility of using input levels seen in circulating tumour DNA (ctDNA) which are typically 1 ng/ml plasma in most cancers. This would allow derivation of blood TMB (28), which has been shown to be a better biomarker of response in PD-1/PD-L1 inhibitor therapy. The assay is also performed at sufficiently high read depth to allow calculation of clonal TMB (29, 30), another marker associated with more accurate identification of potential response to immunotherapy.

In terms of identifying druggable mutations for targeted therapy selection, the TSO pipeline presents an attractive platform especially when coupled with a clinical annotation engine such as the two used here (OncoKDM and PierianDx CGW (31)). We found good correlation between mutations detected in whole genome sequencing experiments, and the identification of druggable mutations was made straightforward by the use of integrated clinical pipelines to produce reproducible data.

Copy number variations, especially amplifications, represent important therapeutic targets. The TSO500 assay detected the known amplifications in a control sample meaning that patients can potentially undergo therapeutic targeting. A unique advantage of the TSO500 system is the ability of a partner targeted RNA-seq assay that can detect RNA fusions. We found that the assay reliably detected NTRK (32), ALK (33), and RET (34) fusions that had previously been identified by FISH, as well as a novel fusion not previously detected using other technologies. Intriguingly, we also successfully detected known fusions at the DNA level de novo in the HD753 control sample, suggesting that this methodology may also be valid for future use, although DNA-based fusion calling has a high false negative rate. Fusion genes represent good drug targets, and a number of novel agents (19, 32) have been shown to be active against fusion genes. Detection of circulating RNA for these fusion genes may also be possible (35) using this assay and could be explored further.

The UK 100,000 Genome project has recently completed, and analysis and reporting is ongoing. The use of whole genome sequencing for tumour-normal pairs using fresh frozen material still has significant challenges from a cost perspective as well as the practicalities of obtaining fresh-frozen tissue over readily available paraffin embedded material. The TSO500 assay costs approximately one third the price of a whole genome sequencing assay, requires no germline DNA control, allows RNA fusion detection, and can be implemented on benchtop sequencers. Its main disadvantages include more laborious library preparation and enrichment chemistry that is vulnerable to drop out.

In conclusion, we believe that the TruSight Oncology 500 assay offers a cost-effective, accurate, pan-cancer assay that can derive SNP, CNV, and gene fusion information across the majority of cancers using a standardised pipeline and therefore is suitable for routine use in precision oncology as a comprehensive genomic profiling solution.

## Data Availability

Data will be made publicly available on EGA on acceptance of the manuscript

## Acknowledgments

We thank Professor Ian Tomlinson (University of Edinburgh) for the contribution of oesophageal cancer DNA samples for this assay.

